# COVID-19 Outcomes Among Users of CD20 Inhibitors for Immune-Mediated Diseases: A Comparative Cohort Study

**DOI:** 10.1101/2021.08.05.21261643

**Authors:** Naomi J. Patel, Kristin M. D’Silva, Tiffany Y-T. Hsu, Michael DiIorio, Xiaoqing Fu, Claire Cook, Lauren Prisco, Lily Martin, Kathleen M.M. Vanni, Alessandra Zaccardelli, Yuqing Zhang, Jeffrey A. Sparks, Zachary S. Wallace

## Abstract

**Objective:** Patients with immune-mediated diseases treated with CD20 inhibitors may have worse COVID-19 outcomes due to impaired humoral immunity, but differences versus the general population are unknown.

**Methods:** We identified patients with immune-mediated diseases who received CD20 inhibitors within one year prior to the index date of PCR-confirmed COVID-19 between January 31, 2020, and January 31, 2021. Comparators with COVID-19 were matched up to 5:1 by age, sex, and PCR date. Hazard ratios (HRs) and 95% confidence intervals (CIs) for hospitalization, mechanical ventilation, and death in CD20 inhibitor users versus comparators were estimated using Cox regression.

**Results:** We identified 114 cases with COVID-19 who had received CD20 inhibitors for immune-mediated diseases (mean age 55 years, 70% female) and 559 matched comparators with COVID-19 (mean age 54 years, 70% female). CD20 inhibitor-treated cases had higher mortality (aHR 2.16; 95% CI: 1.03 to 4.54) than matched comparators. Risks of hospitalization (aHR 0.88; 95% CI: 0.62 to 1.26) and mechanical ventilation (aHR 0.82; 95% CI: 0.36 to 1.87) were similar. Similar trends were seen in analyses according to type of indication (e.g., rheumatic or neurologic disease) and duration of CD20 inhibitor use (<1 or ≥1 year), and after excluding patients with interstitial lung disease, cancer, and those on glucocorticoids prior to COVID-19 diagnosis.

**Conclusions:** Patients who received CD20 inhibitors for immune-mediated diseases prior to COVID-19 had higher mortality following COVID-19 than matched comparators, highlighting the urgent need to mitigate excess risks in CD20 inhibitor users during the ongoing pandemic.

**Key Messages:** *What is already known about this subject?:*

- Patients with immune-mediated diseases treated with CD20 inhibitors may have worse COVID-19 outcomes than those treated with other immunomodulatory medications, but differences compared to the general population are unknown.

*What does this study add?:*

- CD20 inhibitor-treated cases had over two-fold higher risk of mortality than matched general population comparators, although risks of hospitalization and mechanical ventilation were similar.

*How might this impact on clinical practice or future developments?:*

- There is an urgent need for risk mitigation strategies, such as SARS-CoV-2 monoclonal antibodies or booster vaccinations, for patients with immune-mediated diseases treated with CD20 inhibitors during the ongoing COVID-19 pandemic.

## Introduction

Risk of severe coronavirus disease 2019 (COVID-19) outcomes may vary among patients with immune-mediated diseases as a result of disease activity, use of immunosuppressive medications, and comorbid conditions (1-6). Anti-CD20 monoclonal antibodies such as rituximab and ocrelizumab are used to treat immune-mediated autoimmune diseases including rheumatoid arthritis, antineutrophil cytoplasmic antibody (ANCA)-associated vasculitis, and multiple sclerosis, among other conditions. CD20 inhibitors cause elimination of circulating pre-B and B-cells, which results in an impaired immune response to COVID-19 (7, 8).

Results from multiple disease-specific voluntary registries have shown that, compared to patients with immune-mediated diseases on other immunosuppressive or immunomodulatory medications, patients receiving CD20 inhibitors have increased odds of severe COVID-19 outcomes and death (5, 9-11). The risk of severe disease may be particularly high in patients who receive these therapies shortly before contracting COVID-19 (10). These registry data likely impact clinical practice but have limitations, including reporting bias and missing data regarding details of CD20 inhibitor use (e.g., duration of exposure). Additionally, while comparisons of CD20 inhibitor users to patients on other immunosuppressive treatments can address confounding by indication, it is unclear how the treatments used as the reference group in these studies impact the risk of poor COVID-19 outcomes compared to the general population. Therefore, interpretations of estimates generated from prior studies are limited, and the risk of severe COVID-19 outcomes among patients with immune-mediated diseases treated with CD20 inhibitors compared to the general population is poorly understood.

Delays in CD20 inhibitor treatment to minimize COVID-19 risk put patients at risk for disease flare, irreversible organ damage, and disease-related death in some cases. Thus, additional data regarding COVID-19 risks among patients with immune-mediated diseases treated with CD20 inhibitors are needed to inform management decisions during the ongoing pandemic. Here, we evaluate the risk of severe COVID-19 outcomes in patients with immune-mediated diseases treated with CD20 inhibitors versus general population comparators.

## Methods

### Study population

Mass General Brigham (MGB) is a large, multi-center healthcare system with 14 hospitals, including two tertiary care hospitals (Massachusetts General Hospital and Brigham and Women’s Hospital), and multiple primary and specialty outpatient centers in the greater Boston, Massachusetts, area. Using the MGB centralized data warehouse Research Patient Data Registry (RPDR)(12), we identified patients seen at MGB who were ≥18 years of age and had a positive electronic health record (EHR) flag for SARS-CoV-2 (determined by positive molecular testing at MGB or externally, as documented by infection control) between January 31, 2020, and January 31, 2021, and had received a CD20 inhibitor (rituximab, ocrelizumab, ofatumumab, or obinutuzumab) within one year prior to the date of the first positive COVID-19 test (index date). We manually reviewed the EHR to confirm COVID-19 diagnosis and receipt of a CD20 inhibitor prior to COVID-19 diagnosis. Because the RPDR screen only identified patients who received CD20 inhibitors within MGB, we also included patients from our physician-reported cohort of patients with rheumatic disease and confirmed COVID-19 infection, which we have collected from MGB rheumatologists since March 2020 (**Figure 1**). Some of the included cases have been included in prior studies but the observations made in this analysis in comparison to the general population are novel and have not been previously reported (1, 2, 4, 5). Given that our study was focused on patients with immune-mediated diseases, we excluded patients who received CD20 inhibitors for indications related to malignancy or organ transplantation. This study was approved by the MGB Institutional Review Board (2020P000833). Patients were not involved in the design, conduct, or reporting of this study.

**Figure 1.**
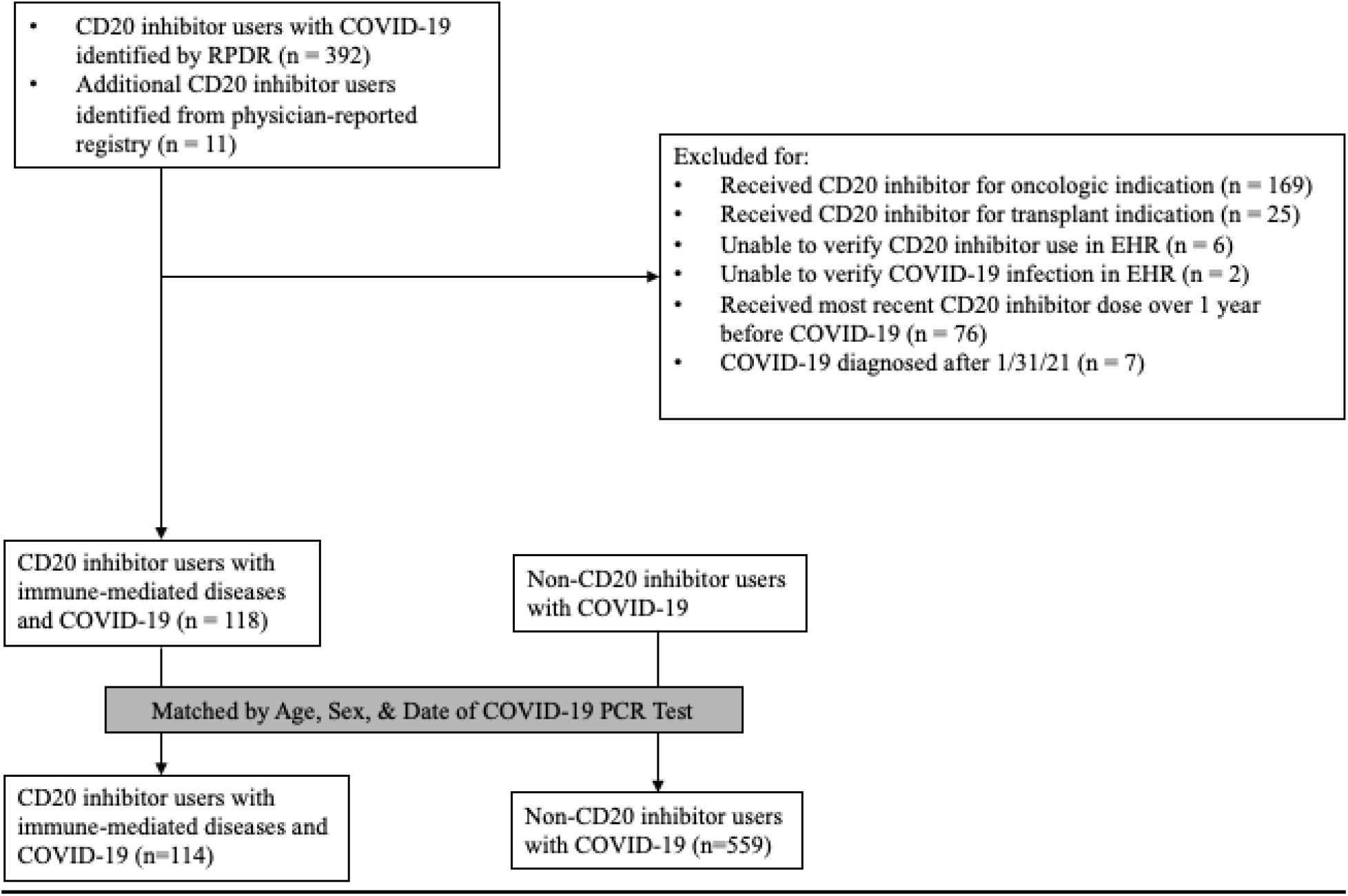
Identification of CD20 inhibitor users with COVID-19. COVID-19, Coronavirus Disease 2019; RPDR, Research Patient Data Repository; PCR, Polymerase Chain Reaction, EHR, Electronic Health Record

### Comparator Identification

Each person with an immune-mediated disease treated with a CD20 inhibitor was matched to up to 5 comparators who had not received CD20 inhibitors from the same COVID-19-positive MGB population, based on age (± 5 years), sex, and the index date (± 5 days). We matched by date of COVID-19 diagnosis because testing criteria and treatment strategies changed over time.

### Covariates

For the CD20 inhibitor users, clinical variables of interest were extracted from the EHR by manual EHR review, including the indication for the CD20 inhibitor and dates of initial and most recent CD20 inhibitor administration. Other covariates of interest including immune-mediated disease diagnosis and duration, concomitant immunomodulatory medications (including specific dose of any glucocorticoid when available), and disease activity level (based on global assessment by the treating provider as documented in the EHR) were also obtained from EHR review.

For CD20 inhibitor users and comparators, additional variables were extracted from the COVID-19 Data Mart (13), an EHR-based data enclave established by MGB that includes all patients diagnosed with COVID-19. Covariates extracted from the COVID-19 Data Mart included demographics (age, sex, and self-identified race/ethnicity), smoking status, and medical comorbidities. Baseline characteristics including demographics, comorbidities, smoking history, and body mass index (BMI) were assessed in the one year prior to the index date, and the Charlson Comorbidity Index (CCI) (14) was calculated using all available data prior to the index date.

### Outcome Assessment

Mortality, the primary outcome, was ascertained from the Data Mart but also confirmed by manual EHR review and online searches of obituaries for all cases and comparators to capture deaths that may have occurred outside of the system (15). Secondary outcomes extracted from the Data Mart included hospitalization and mechanical ventilation.

### Subgroup and Sensitivity Analyses

We performed several subgroup analyses in which we limited cases to those with rheumatic disease or neurologic indications (separate analyses), recent CD20 exposure (within 3 months of index date), short-term duration of CD20 inhibitor exposure (<1 year), and long-term duration of CD20 inhibitor exposure (≥1 year). In sensitivity analyses, we excluded patients with interstitial lung disease or cancer (separate analyses) since these are indications for CD20 inhibitor use in some rheumatic diseases and may be independently associated with COVID-19 severity. We also performed a sensitivity analysis in which we excluded patients who also used glucocorticoids at the time of COVID-19 infection since this may be independently associated with COVID-19 severity. In these subgroup and sensitivity analyses, included cases were compared to their general population comparators.

### Statistical Analysis

Categorical variables are presented as number (percentage), and continuous variables are presented as mean ± standard deviation or median ± interquartile range, as appropriate. Continuous variables were compared using a two-sample *t*-test for continuous normally distributed variables or Wilcoxon test for continuous non-normally distributed variables. Categorical variables were compared using Chi-square tests.

The index date was the date of COVID-19 diagnosis by molecular testing. Person-days of follow-up were determined for each subject from the index date to the first of the following: occurrence of the outcome of interest, date of the last encounter at MGB, or end of the study period (3/2/2021, to allow at least 30 days of follow-up per person). We calculated incidence rates per 1000 days by dividing the number of events by the number of person-days. Multivariable Cox proportional hazard regression models were used to estimate hazard ratios (HR) and 95% confidence intervals (CI) for hospitalization, mechanical ventilation, and death in separate models, comparing CD20 inhibitor users to non-users. The first multivariable model adjusted for age. The second multivariable model adjusted for age and race. The third multivariable model (the fully-adjusted model) adjusted for age, race, BMI, and CCI (dichotomized as < 2 or ≥2). Because of a limited number of observed outcomes in some subgroup and sensitivity analyses, only unadjusted or partially adjusted models are reported in some instances (i.e., when <7 outcomes per covariate were present, adjusted models were not performed). For analyses of risk of hospitalization and mechanical ventilation, death was treated as a competing risk using a cause-specific model yielding subdistribution HRs (16). The level of significance was set as a two-tailed p<0.05, and statistical analyses were completed using SAS statistical software (version 9.4; SAS Institute, Inc.).

## Results

We identified 114 patients with COVID-19 who had an immune-mediated disease treated with a CD20 inhibitor within 12 months preceding COVID-19 diagnosis for a non-oncologic and non-transplant indication and 559 matched comparators. The mean age was 55 years in the CD20 inhibitor group and 54 years in the comparator group, and 70% were female in each group (**Table 1**). The distribution of race/ethnicity was similar between groups. The Charlson Comorbidity Index was higher in the CD20 inhibitor group (median of 1 vs. 0, p=0.001).

**Table 1.** Clinical characteristics of immune-mediated cases with CD20 inhibitor use prior to COVID-19 and age, sex, and COVID-19 diagnosis date-matched comparators.

| **Characteristic** | **CD20 Inhibitor-Treated Immune-Mediated Cases (n=114)** | **Matched Comparators (n=559)** | **p-value** |
| --- | --- | --- | --- |
| Age, years, mean ± SD | 55 ± 15 | 54 ± 15 | 0.44 |
| Female, n (%) | 80 (70) | 391 (70) | 0.96 |
| Race, n (%) |  |  |  |
| White | 69 (61) | 316 (57) | 0.43 |
| Black or African American | 16 (14) | 70 (13) |  |
| Asian | 4 (4) | 12 (2) |  |
| Other | 25 (22) | 161 (29) |  |
| Hispanic or Latinx ethnicity, n (%) | 6 (5) | 46 (8) | 0.28 |
| Body mass index, kg/m<sup>2</sup>, mean ± SD | 28.7 ± 6.2 | 29.7 ± 6.7 | 0.16 |
| Smoking status, n (%) |  |  |  |
| Never | 66 (58) | 271 (48) | 0.06 |
| Former | 29 (25) | 126 (23) |  |
| Current | 3 (3) | 18 (3) |  |
| Unknown | 16 (14) | 144 (26) |  |
| Charlson Comorbidity Index, median (IQR) | 1 (0, 2) | 0 (0, 1) | 0.001 |
| Comorbidities, n (%) |  |  |  |
| Hypertension | 45 (39) | 120 (21) | <0.0001 |
| Diabetes | 13 (11) | 56 (10) | 0.66 |
| Coronary artery disease | 8 (7) | 21 (4) | 0.12 |
| Heart failure | 8 (7) | 11 (2) | 0.003 |
| Asthma | 10 (9) | 32 (6) | 0.22 |
| Chronic obstructive pulmonary disease | 6 (5) | 6 (1) | 0.002 |
| Obstructive sleep apnea | 2 (2) | 25 (4) | 0.18 |
| Chronic kidney disease | 12 (11) | 26 (5) | 0.01 |
| Interstitial lung disease | 25 (22) | 55 (10) | < 0.001 |
| Malignancy | 11 (10) | 6 (1) | < 0.001 |

COVID-19, Coronavirus Disease 2019; SD, standard deviation; IQR, interquartile range.

Among the immune-mediated disease patients who received a CD20 inhibitor, 90 (79%) received rituximab and 26 (23%) received ocrelizumab; two patients had received rituximab initially but were later switched to ocrelizumab (**Table 2**). The most common indication for CD20 inhibitor use was rheumatic disease (54 [47%]), followed by neurologic conditions (43 [38%]) (**Table 2**). The duration of CD20 inhibitor use was <1 year in 33 (29%) patients, 1-3 years in 51 (45%), and >3 years in 30 (26%). Forty-eight patients (42%) had received their most recent CD20 inhibitor infusion within three months of COVID-19 diagnosis.

**Table 2.** Immune-mediated disease characteristics of cases with CD20 inhibitor use prior to COVID-19.

| **Characteristic** | **CD20 Inhibitor-Treated Immune-Mediated Cases (n=114)** |
| --- | --- |
| Indication for CD20 inhibitor, n (%) |  |
| Rheumatic condition only | 54 (47) |
| Inflammatory arthritis* | 19 (17) |
| Vasculitis | 17 (15) |
| Systemic lupus erythematosus | 7 (6) |
| Inflammatory myositis | 6 (5) |
| Other rheumatic condition† | 3 (3) |
| Multiple primary rheumatic diagnoses‡ | 2 (2) |
| Neurologic condition | 43 (38) |
| Multiple sclerosis | 33 (29) |
| Neuromyelitis optica | 4 (4) |
| Other neurologic condition§ | 6 (5) |
| Ocular inflammation | 3 (3) |
| Hematologic condition only | 5 (4) |
| Thrombotic thrombocytopenic purpura | 3 (3) |
| Autoimmune hemolytic anemia | 2 (2) |
| Autoimmune hepatitis | 3 (3) |
| Both rheumatic and hematologic conditions\|\| | 2 (2) |
| Other miscellaneous conditions¶ | 4 (4) |
| Immune-mediated disease duration, years, median (IQR) | 6 (3, 15) |
| Immune-mediated disease status |  |
| Remission | 26 (23) |
| Low activity | 58 (51) |
| Moderate/high activity | 30 (26) |
| Type of CD20 inhibitor, n (%)** |  |
| Rituximab or biosimilar | 90 (79) |
| Ocrelizumab | 26 (23) |
| Duration of CD20 inhibitor use, n (%) |  |
| <1 year | 33 (29) |
| 1-3 years | 51 (45) |
| >3 years | 30 (26) |
| Most recent CD20 inhibitor dose prior to COVID-19 onset, n (%) |  |
| <3 months | 48 (42) |
| 3-6 months | 44 (39) |
| 6-12 months | 22 (19) |
| Concomitant immunomodulatory medications at COVID-19 onset, n (%) |  |
| Mycophenolate mofetil | 8 (7) |
| Methotrexate | 6 (5) |
| Hydroxychloroquine | 5 (4) |
| Leflunomide | 3 (3) |
| Other immunomodulatory medication†† | 6 (5) |
| Oral glucocorticoid | 35 (31) |
| Prednisone-equivalent daily dose mg, median (IQR) | 7.5 (5.0, 15.0) |

\* Includes rheumatoid arthritis, other inflammatory arthritis, and juvenile idiopathic arthritis.
† Includes Sjögren's syndrome and IgG4-related disease.
‡ Includes one patient with both rheumatoid arthritis and vasculitis and one patient with both inflammatory myopathy and inflammatory arthritis
§ Includes myasthenia gravis, autoimmune encephalitis, acute disseminated encephalomyelitis, cavernous sinus mass, and small fiber polyneuropathy.
|| Includes a patient with rheumatoid arthritis and immune thrombocytopenic purpura (ITP), and a patient with vasculitis and antiphospholipid syndrome
¶ Includes pemphigus vulgaris, 2 membranous nephropathy, and autoimmune interstitial lung disease.
\*\* No individuals were on obinutuzumab or ofatumumab. Two individuals were initially on rituximab and then transitioned to ocrelizumab.
†† Includes azathioprine (n=1), cyclophosphamide (n=2), sulfasalazine (n=1), and tacrolimus (n=2).

Patients with immune-mediated disease treated with CD20 inhibitors had a higher risk of death (12 [11%] vs. 21 [4%]; adjusted HR 2.16; 95% CI: 1.03 to 4.54) than matched comparators. The risks of hospitalization (35 [31%] vs. 123 [22%]; adjusted HR 0.88; 95% CI: 0.62 to 1.26) and mechanical ventilation (6 [5%] vs. 26 [5%]; adjusted HR 0.82; 95% CI: 0.36 to 1.87) were similar in both groups (**Table 3**). Of note, in both groups, more patients died than were mechanically ventilated (**Supplemental Table 1**). Among CD20 inhibitor users, 6 (5%##) had a code status that indicated “Do Not Intubate” and ultimately died compared with 10 (2##%) in the comparator group. These differences suggest that the risk of needing mechanical ventilation among CD20 inhibitor users is likely an underestimate. No deaths were noted to be directly related to the immune-mediated disease. Among immune-mediated disease patients on CD20 inhibitors who developed COVID-19, 9 patients (8%) had COVID-19 antibodies tested after COVID-19 diagnosis (median 70 days, IQR 35 to 87 days) for clinical indications. Of these 9 patients, 2 had positive COVID-19 antibodies, and 7 patients had negative COVID-19 antibodies.

**Table 3.**
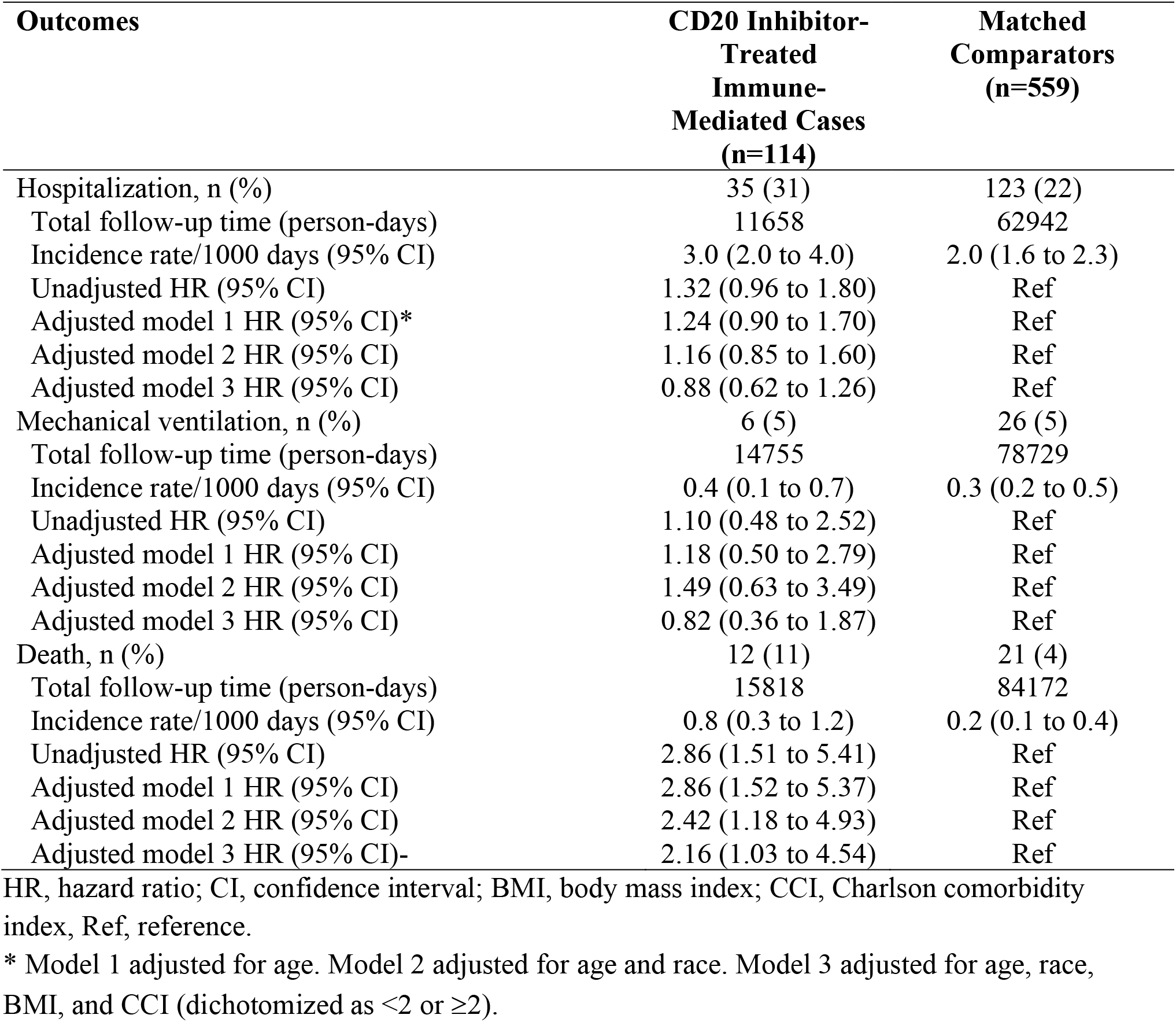
COVID-19 outcomes in immune-mediated cases treated with CD20 inhibitors versus matched comparators.

| **Outcomes** | **CD20 Inhibitor-Treated Immune-Mediated Cases (n=114)** | **Matched Comparators (n=559)** |
| --- | --- | --- |
| Hospitalization, n (%) | 35 (31) | 123 (22) |
| Total follow-up time (person-days) | 11658 | 62942 |
| Incidence rate/1000 days (95% CI) | 3.0 (2.0 to 4.0) | 2.0 (1.6 to 2.3) |
| Unadjusted HR (95% CI) | 1.32 (0.96 to 1.80) | Ref |
| Adjusted model 1 HR (95% CI)* | 1.24 (0.90 to 1.70) | Ref |
| Adjusted model 2 HR (95% CI) | 1.16 (0.85 to 1.60) | Ref |
| Adjusted model 3 HR (95% CI) | 0.88 (0.62 to 1.26) | Ref |
| Mechanical ventilation, n (%) | 6 (5) | 26 (5) |
| Total follow-up time (person-days) | 14755 | 78729 |
| Incidence rate/1000 days (95% CI) | 0.4 (0.1 to 0.7) | 0.3 (0.2 to 0.5) |
| Unadjusted HR (95% CI) | 1.10 (0.48 to 2.52) | Ref |
| Adjusted model 1 HR (95% CI) | 1.18 (0.50 to 2.79) | Ref |
| Adjusted model 2 HR (95% CI) | 1.49 (0.63 to 3.49) | Ref |
| Adjusted model 3 HR (95% CI) | 0.82 (0.36 to 1.87) | Ref |
| Death, n (%) | 12 (11) | 21 (4) |
| Total follow-up time (person-days) | 15818 | 84172 |
| Incidence rate/1000 days (95% CI) | 0.8 (0.3 to 1.2) | 0.2 (0.1 to 0.4) |
| Unadjusted HR (95% CI) | 2.86 (1.51 to 5.41) | Ref |
| Adjusted model 1 HR (95% CI) | 2.86 (1.52 to 5.37) | Ref |
| Adjusted model 2 HR (95% CI) | 2.42 (1.18 to 4.93) | Ref |
| Adjusted model 3 HR (95% CI)- | 2.16 (1.03 to 4.54) | Ref |

HR, hazard ratio; CI, confidence interval; BMI, body mass index; CCI, Charlson comorbidity index, Ref, reference.
\* Model 1 adjusted for age. Model 2 adjusted for age and race. Model 3 adjusted for age, race, BMI, and CCI (dichotomized as <2 or ≥2).

In subgroup analyses, we found that there was a similar trend among patients with rheumatic disease (**Table 4**) as the indication for CD20 inhibitor use. We found numerically higher risk of death among patients with neurologic disease as the indication for CD20 inhibitor use, though analysis was limited by low number of events (**Table 5**). Similar trends were also observed among short- and long-term CD20 inhibitor users, those with recent (within 3 months) exposure to a CD20 inhibitor, and after excluding patients with interstitial lung disease, cancer, and those receiving glucocorticoids prior to COVID-19 diagnosis (**Supplemental Tables 2-6**). Our ability to detect significant differences and perform multivariable adjustment in these subgroup analyses was limited by the number of observed events.

**Table 4.**
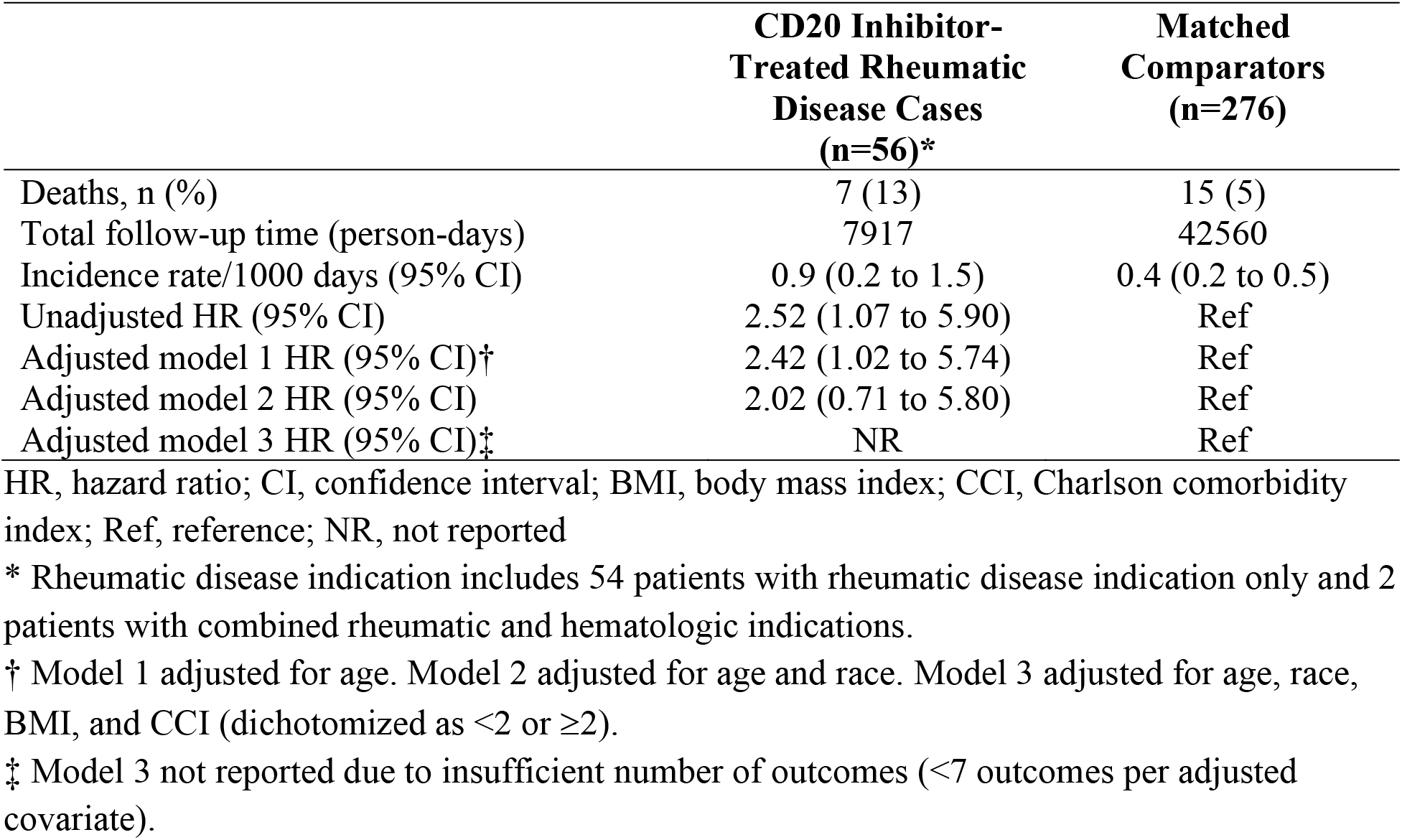
Risk of death following COVID-19 in patients treated with CD20 inhibitors for rheumatic disease indications versus comparators.

**Table 5.**
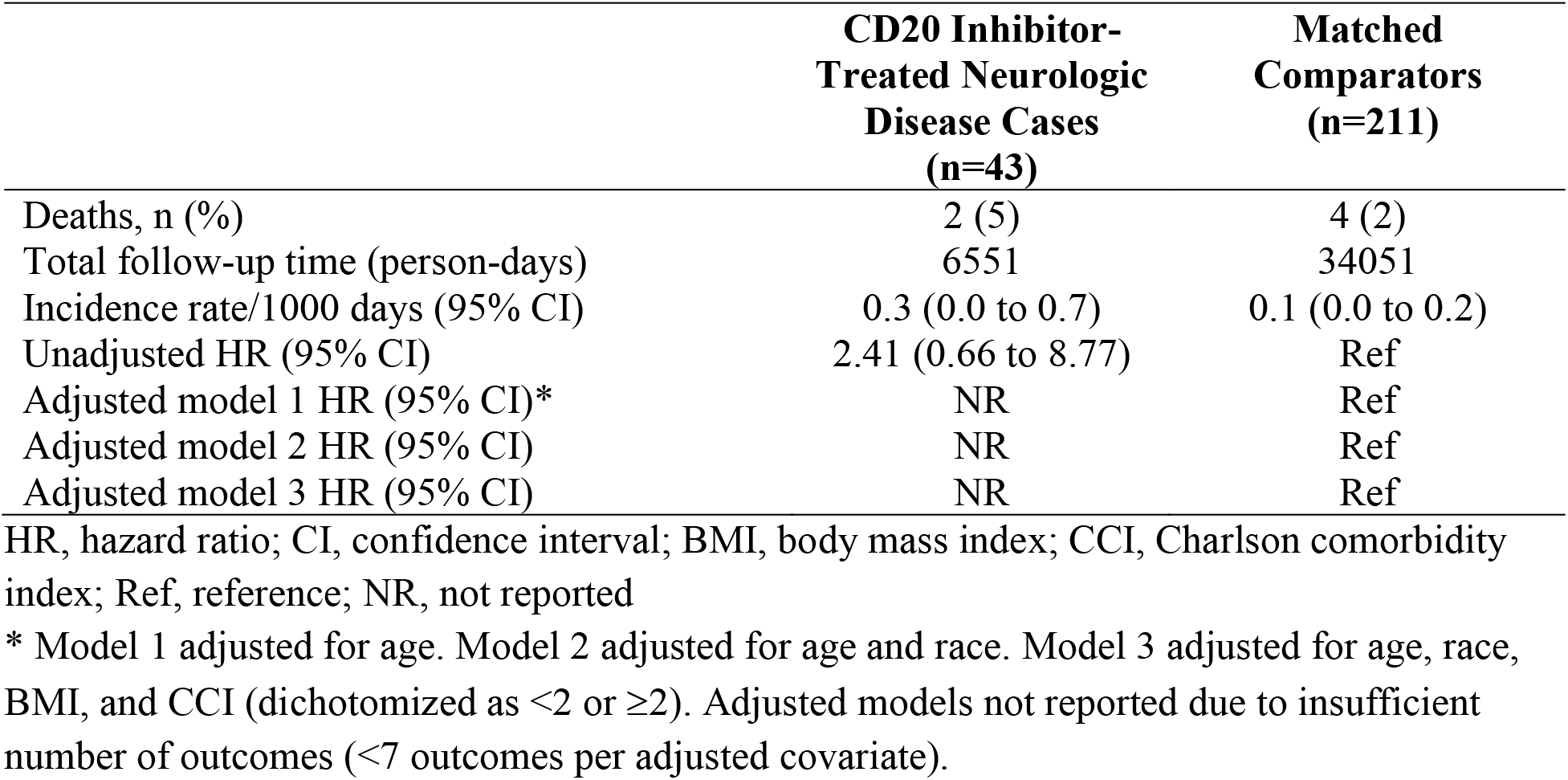
Risk of death following COVID-19 in patients treated with CD20 inhibitors for neurologic disease indications versus comparators.

## Discussion

In this multicenter cohort study, we found an increased risk of death following COVID-19 infection among patients with immune-mediated disease treated with CD20 inhibitors compared to general population comparators. This association was observed among more recent CD20 inhibitor initiators as well as among patients who had been on long-standing CD20 inhibitor treatment. The results remained consistent in analyses adjusting for comorbidity burden and across sensitivity and subgroup analyses meant to further address potential confounders. These findings raise concern regarding the impact of CD20 inhibitor exposure on COVID-19 risk and the need to investigate strategies to mitigate this potential risk, especially given the poor response to vaccines observed among CD20 inhibitor users (17, 18).

CD20 inhibitors have been a particular concern during the COVID-19 pandemic because the humoral immune response plays an important role in the response to SARS-CoV-2 infection (8, 19, 20). During the early phase of infection, high antibody titers are associated with higher levels of neutralizing antibodies to the receptor binding domain of the spike protein, and antibody titers are significantly higher in patients with shorter duration of SARS-CoV-2 RNA positivity (21, 22). To this end, there have been several reports of patients receiving CD20 inhibitors with subsequent prolonged courses of COVID-19 (22-26). Post-mortem studies of lymph nodes from patients with fatal COVID-19 showed the absence of germinal centers and a reduction in germinal center B and T cells, suggesting that a dysregulated adaptive immune response occurs in fatal COVID-19 (27). Lastly, patients treated with CD20 inhibitors had a 36-fold reduction in humoral responses compared to immunocompetent patients at 1-2 weeks after the second dose of SARS-CoV-2 mRNA vaccines (17, 18, 28). Further research is needed to determine if prophylactic and therapeutic strategies such as convalescent plasma or SARS-CoV-2 monoclonal antibody therapy may be beneficial in the prevention and treatment of COVID-19 in patients with B cell depletion (29, 30).

In light of the critical importance of antibody formation for a robust immune response to COVID-19, we had hypothesized that long-term CD20 inhibitor use would be associated with a greater risk of death compared to the general population because of more sustained B cell depletion. Instead, we found a similar risk among short-term (<1 year) and long-term (≥1 year) CD20 inhibitor users. It is possible that long-term CD20 inhibitor users are a generally healthier cohort who have previously tolerated CD20 inhibition well without infections or other complications, thus resulting in similar risk of severe COVID-19 compared to short-term CD20 inhibitor users. However, our observations may suggest that continuous B cell depletion versus more recent B cell depletion initiation may not have a differential impact on the critical immediate immunologic response that is blunted by CD20 inhibitors and needed to control infection (31, 32). We cannot rule out the possibility that small sample sizes may have limited the ability to detect a difference when comparing outcomes among short-term and long-term CD20 inhibitor users. Similar to Avouac *et al*., we also found a significantly higher risk of death among those who had received their most recent CD20 inhibitor close to the time of COVID-19 infection (10).

Despite the increased risk of death, we did not find any statistical difference in risk of hospitalization or mechanical ventilation between those who received CD20 inhibitors and matched comparators. This discrepancy between increased risk of death but not mechanical ventilation may be explained by the use of “Do Not Intubate” orders. These orders may have led to an underestimate of the effect of CD20 inhibitor use on mechanical ventilation risk if the proportion needing (but not receiving) mechanical ventilation was actually greater among the CD20 inhibitor users, as our findings suggest. We further conducted subgroup analyses to better understand specific factors driving the increased risk of death with CD20 inhibitor exposure and found that the risk was attenuated in multivariable models but similar trends persisted despite low event rates. Indeed, our findings persisted when we excluded patients with interstitial lung disease or malignancy, two populations commonly prescribed rituximab for immune-mediated conditions who might be at particularly high risk for poor COVID-19 outcomes because of other medical comorbidities.

Our study has multiple strengths. First, we systematically identified patients who received CD20 inhibitors for a variety of immune-mediated conditions in a large healthcare system, increasing the generalizability of our observations to the diverse populations who use CD20 inhibitors. Second, details regarding CD20 inhibitor indication, length of exposure, and COVID-19 outcomes were available, in contrast to prior registry-based studies.

Despite these strengths, our study has certain limitations. First, though our findings persisted after adjustment for covariates, there may be residual confounding by CD20 inhibitor indication, concomitant glucocorticoid use, or disease severity, as CD20 inhibitors are often used as initial induction therapy for severe immune-mediated diseases (e.g., ANCA-associated vasculitis) or as treatment for diseases that have been refractory to other therapies (e.g., rheumatoid arthritis). However, our findings remained robust in sensitivity analyses and subgroup analyses that addressed the potential impact of residual confounding. Regardless, one should cautiously interpret these results as applying to the population of patients with immune-mediated diseases treated with CD20 inhibitors in the context of the known effects of CD20 inhibitors on B cell responses and antibody production. Second, multivariable adjustment was limited in some subgroup and sensitivity analyses due to low event rates. However, the observed trends remained consistent with those observed in the primary analysis.

In conclusion, we found an increased risk of death in patients with immune-mediated diseases who had received CD20 inhibitors prior to COVID-19 diagnosis. CD20 inhibitors are the standard of care for induction and maintenance treatment of multiple immune-mediated diseases, some of which have few alternatives. Additional studies are needed to evaluate the potential use of anti-SARS-CoV-2 monoclonal antibodies, booster vaccinations, and other strategies to reduce the risk of poor COVID-19 outcomes in this population. Providers should interpret these results cautiously and weigh the risks and benefits of ongoing CD20 inhibitor use on an individual basis using shared decision-making.

## Data Availability

Data are available upon request.

## Acknowledgements

None

