## Supplement for "COVID-19 Outcomes Among Users of CD20 Inhibitors for Immune-Mediated Diseases: A Comparative Cohort Study"

**Supplemental Table 1. Details of death following COVID-19 in immune-mediated cases treated with CD20 inhibitors and matched comparators.**

| | CD20 Inhibitor-Treated Immune-Mediated Case Deaths (n=12) | Matched Comparator Deaths (n=21) |
|---|---|---|
| Age, years, mean ± SD | 69.0 ± 19.4 | 77.0 ± 11.7 |
| Female, n (%) | 5 (42) | 6 (29) |
| Hospitalized within MGB, n (%) | 11 (92) | 20 (95) |
| Mechanically ventilated within MGB, n (%) | 4 (33) | 8 (38) |
| "Do not intubate" code status, n (%) | 6 (50) | 10 (48) |

MGB, Mass General Brigham healthcare system; SD, standard deviation; COVID-19, Coronavirus Disease 2019.

**Supplemental Table 2. Risk of death following COVID-19 in patients treated with CD20 inhibitors for rheumatic disease indications versus comparators.**

| | **CD20 Inhibitor-Treated Rheumatic Disease Cases (n=56)*** | **Matched Comparators (n=276)** |
|---|---|---|
| Deaths, n (%) | 7 (13) | 15 (5) |
| Total follow-up time (person-days) | 7917 | 42560 |
| Incidence rate/1000 days (95% CI) | 0.9 (0.2 to 1.5) | 0.4 (0.2 to 0.5) |
| Unadjusted HR (95% CI) | 2.52 (1.07 to 5.90) | Ref |
| Adjusted model 1 HR (95% CI)† | 2.42 (1.02 to 5.74) | Ref |
| Adjusted model 2 HR (95% CI) | 2.02 (0.71 to 5.80) | Ref |
| Adjusted model 3 HR (95% CI)‡ | NR | Ref |

HR, hazard ratio; CI, confidence interval; BMI, body mass index; CCI, Charlson comorbidity index; Ref, reference; NR, not reported

* Rheumatic disease indication includes 54 patients with rheumatic disease indication only and 2 patients with combined rheumatic and hematologic indications.

† Model 1 adjusted for age. Model 2 adjusted for age and race. Model 3 adjusted for age, race, BMI, and CCI (dichotomized as <2 or ≥2).

‡ Model 3 not reported due to insufficient number of outcomes (<7 outcomes per adjusted covariate).

**Supplemental Table 3. Risk of death following COVID-19 in patients treated with CD20 inhibitors for neurologic disease indications versus comparators.**

| | CD20 Inhibitor-Treated Neurologic Disease Cases (n=43) | Matched Comparators (n=211) |
|---|---|---|
| Deaths, n (%) | 2 (5) | 4 (2) |
| Total follow-up time (person-days) | 6551 | 34051 |
| Incidence rate/1000 days (95% CI) | 0.3 (0.0 to 0.7) | 0.1 (0.0 to 0.2) |
| Unadjusted HR (95% CI) | 2.41 (0.66 to 8.77) | Ref |
| Adjusted model 1 HR (95% CI)* | NR | Ref |
| Adjusted model 2 HR (95% CI) | NR | Ref |
| Adjusted model 3 HR (95% CI) | NR | Ref |

HR, hazard ratio; CI, confidence interval; BMI, body mass index; CCI, Charlson comorbidity index; Ref, reference; NR, not reported

* Model 1 adjusted for age. Model 2 adjusted for age and race. Model 3 adjusted for age, race, BMI, and CCI (dichotomized as <2 or ≥2). Adjusted models not reported due to insufficient number of outcomes (<7 outcomes per adjusted covariate).

**Supplemental Table 4. Risk of death following COVID-19 in immune-mediated cases treated with CD20 inhibitors for greater than or less than 1 year versus matched comparators.**

| | < 1 Year of CD20 Inhibitor Exposure | | > 1 Year of CD20 Inhibitor Exposure | |
|---|---|---|---|---|
| | CD20 Inhibitor-Treated Immune-Mediated Cases (n=81) | Matched Comparators (n=399) | CD20 Inhibitor-Treated Immune-Mediated Cases (n=33) | Matched Comparators (n=160) |
| Deaths, n (%) | 7 (9) | 11 (3) | 5 (15) | 10 (6) |
| Total follow-up time (person-days) | 11088 | 59458 | 4730 | 24714 |
| Incidence rate/1000 days (95% CI) | 0.6 (0.2 to 1.1) | 0.2 (0.1 to 0.3) | 1.1 (0.1 to 2.0) | 0.4 (0.2 to 0.7) |
| Unadjusted HR (95% CI) | 2.82 (1.34 to 5.96) | Ref | 2.92 (0.95 to 8.99) | Ref |
| Adjusted model 1 HR (95% CI)* | 2.85 (1.35 to 6.01) | Ref | 2.90 (0.95 to 8.87) | Ref |
| Adjusted model 2 HR (95% CI) | 2.91 (1.26 to 6.74) | Ref | 2.02 (0.61 to 6.70) | Ref |
| Adjusted model 3 HR (95% CI)† | NR | Ref | NR | Ref |

HR, hazard ratio; CI, confidence interval; BMI, body mass index; CCI, Charlson comorbidity index; Ref, reference; NR, not reported

* Model 1 adjusted for age. Model 2 adjusted for age and race. Model 3 adjusted for age, race, BMI, and CCI (dichotomized as <2 or ≥2).

† Model 3 not reported due to insufficient number of outcomes (<7 outcomes per adjusted covariate).

**Supplemental Table 5. Risk of death following COVID-19 in patients treated with CD20 inhibitors and not receiving glucocorticoids versus comparators.**

| | CD20 Inhibitor-Treated Cases (n=79) | Matched Comparators (n=384) |
|---|---|---|
| Deaths, n (%) | 6 (8) | 6 (2) |
| Total follow-up time (person-days) | 11878 | 61304 |
| Incidence rate/1000 days (95% CI) | 0.50 (0.10 to 0.90) | 0.10 (0.00 to 0.20) |
| Unadjusted HR (95% CI) | 4.37 (1.79 to 10.71) | Ref |
| Adjusted model 1 HR (95% CI)* | 4.43 (1.79 to 10.98) | Ref |
| Adjusted model 2 HR (95% CI)† | NR | Ref |
| Adjusted model 3 HR (95% CI) | NR | Ref |

HR, hazard ratio; CI, confidence interval; BMI, body mass index; CCI, Charlson comorbidity index; Ref, reference; NR, not reported

* Model 1 adjusted for age. Model 2 adjusted for age and race. Model 3 adjusted for age, race, BMI, and CCI (dichotomized as <2 or ≥2).

† Models 2 and 3 not reported due to insufficient number of outcomes (<7 outcomes per adjusted covariate).

**Supplemental Table 6. Risk of death following COVID-19 in patients without cancer treated with CD20 inhibitors versus comparators.**

| | CD20 Inhibitor-Treated Cases (n=103) | Matched Comparators (n=502) |
|---|---|---|
| Deaths, n (%) | 11 (11) | 19 (4) |
| Total follow-up time (person-days) | 14558 | 76981 |
| Incidence rate/1000 days (95% CI) | 0.80 (0.30 to 1.20) | 0.20 (0.10 to 0.40) |
| Unadjusted HR (95% CI) | 3.00 (1.53 to 5.92) | Ref |
| Adjusted model 1 HR (95% CI)* | 2.99 (1.52 to 5.86) | Ref |
| Adjusted model 2 HR (95% CI) | 2.57 (1.21 to 5.45) | Ref |
| Adjusted model 3 HR (95% CI) | 2.20 (1.00 to 4.85) | Ref |

HR, hazard ratio; CI, confidence interval; BMI, body mass index; CCI, Charlson comorbidity index.

* Model 1 adjusted for age. Model 2 adjusted for age and race. Model 3 adjusted for age, race, BMI, and CCI (dichotomized as <2 or ≥2).

**Supplemental Table 7. Risk of death following COVID-19 in patients without interstitial lung disease treated with CD20 inhibitors versus comparators.**

| | CD20 Inhibitor-Treated Cases (n=89) | Matched Comparators (n=396) |
|---|---|---|
| Deaths, n (%) | 6 (7) | 7 (2) |
| Total follow-up time (person-days) | 13535 | 65014 |
| Incidence rate/1000 days (95% CI) | 0.40 (0.10 to 0.80) | 0.10 (0.00 to 0.20) |
| Unadjusted HR (95% CI) | 3.51 (1.41 to 8.71) | Ref |
| Adjusted model 1 HR (95% CI)* | 3.45 (1.36 to 8.73) | Ref |
| Adjusted model 2 HR (95% CI)† | NR | Ref |
| Adjusted model 3 HR (95% CI) | NR | Ref |

HR, hazard ratio; CI, confidence interval; BMI, body mass index; CCI, Charlson comorbidity index; Ref, reference; NR, not reported
* Model 1 adjusted for age. Model 2 adjusted for age and race. Model 3 adjusted for age, race, BMI, and CCI (dichotomized as <2 or ≥2).
† Models 2 and 3 not reported due to insufficient number of outcomes (<7 outcomes per adjusted covariate).

**Supplemental Table 8. Risk of death following COVID-19 in patients treated with most recent CD20 inhibitor dose within 3 months of COVID-19 infection versus comparators.**

| | CD20 Inhibitor-Treated Cases (n=48) | Matched Comparators (n=235) |
|---|---|---|
| Deaths, n (%) | 7 (15) | 6 (3) |
| Total follow-up time (person-days) | 6099 | 34753 |
| Incidence rate/1000 days (95% CI) | 1.10 (0.30 to 2.00) | 0.20 (0.00 to 0.30) |
| Unadjusted HR (95% CI) | 9.59 (2.98 to 30.85) | Ref |
| Adjusted model 1 HR (95% CI)* | 9.79 (3.00 to 31.93) | Ref |
| Adjusted model 2 HR (95% CI)† | NR | Ref |
| Adjusted model 3 HR (95% CI) | NR | Ref |

HR, hazard ratio; CI, confidence interval; BMI, body mass index; CCI, Charlson comorbidity index; Ref, reference; NR, not reported
* Model 1 adjusted for age. Model 2 adjusted for age and race. Model 3 adjusted for age, race, BMI, and CCI (dichotomized as <2 or ≥2).
† Models 2 and 3 not reported due to insufficient number of outcomes (<7 outcomes per adjusted covariate).

******POTENTIAL Supplemental Table 9. Immune-mediated disease status at the time of COVID-19 diagnosis by timing of most recent rituximab dose prior to infection. ******

| | Most recent CD20 inhibitor dose < 3 months prior to COVID-19 (n=48) | Most recent CD20 inhibitor dose 3-6 months prior to COVID-19 (n=44) | Most recent CD20 inhibitor dose 6-12 months prior to COVID-19 (n=22) | P-value |
|---|---|---|---|---|
| Immune-mediated disease status | | | | 0.23 |
| Remission | 13 (27) | 7 (16) | 6 (27) | |
| Low activity | 19 (40) | 28 (64) | 11 (50) | |
| Moderate/high activity | 16 (33) | 9 (20) | 5 (23) | |

| **Number of death** | **Most recent CD20 inhibitor dose < 3 months prior to COVID-19 (n=48)** | **Most recent CD20 inhibitor dose 3-6 months prior to COVID-19 (n=44)** | **Most recent CD20 inhibitor dose 6-12 months prior to COVID-19 (n=22)** |
|---|---|---|---|
| Immune-mediated disease status | | | |
| Remission | 1 (2) | 1 (2) | 0 (0) |

| | | | |
|---|---|---|---|
| Low activity | 1 (2) | 2 (4) | 0 (0) |
| Moderate/high activity | 5 (10) | 1 (2) | 1 (5) |
